# A student made MOOC for medical students during the SARS-CoV-2 pandemic

**DOI:** 10.1101/2020.07.05.20146654

**Authors:** David Fabian Ramirez Moreno

**Affiliations:** Universidad pedagógica y tecnológica de Colombia (UPTC); Grupo de epidemiología clínica de Colombia (GRECO)

**Keywords:** COVID-19, Quarantine, Epilepsy, Latin America, (Source: MeSH-NLM)

## Abstract

**Background:** Latin America was one of the last regions of the world to become affected by the COVID-19 pandemic. As a response to the emergency virtual education was implemented in almost every country of the region.

**Methods:** a massive open online course about epilepsy was made using only free software and platforms following the international league against epilepsy competence-based domains for epileptology teaching.

**Results:** 250 healthcare students signed up for the course and only 17.2% of them had previously participated in courses like this one. This course had a completion rate of 37.2% and of the students that completed the course 98.3% would participate in course like this in the future.

**Conclusion:** In conclusion MOOCs can be easily implemented as a powerful pedagogic strategy during the COVID-19 pandemic and can have a positive impact not only in in its proposed learning objectives but it can help closing the gap that prevent Latin American healthcare students to acquire actively knowledge trough them.

**Personal, Professional, and Institutional Social Network accounts:**

- **Facebook:** https://www.facebook.com/davidfabianrm
- **Facebook:**https://www.facebook.com/Grupo-de-Investigaci%C3%B3n-en-Epidemiolog%C3%ADa-Cl%C3%ADnica-de-Colombia-GRECO-107921290887449

**Discussion Points:**

1. How can a student made MOOC impact Latin American medical students?
2. Can a student made MOOC encourage medical students to use and discover new educational online offers?
3. COVID-19 pandemic can be a good time to improve the academy using new technologies and pedagogic strategies.
4. MOOCs can become a powerful tool for learning and teaching worldwide during the COVID-19 pandemic.
5. MOOCs are an attractive tool for teaching and learning during the social distancing caused by COVID-19.

## INTRODUCTION

Latin America was one of the last regions of the world to become affected by the COVID-19 pandemic, when it happened its different governments started implementing measures of hygiene, social distancing and quarantine that affected almost every aspect of the lives of their inhabitants. One of these aspects was the education. Every country of the region took action on the matter and the restrictions ranged from the complete suspension of the classroom classes to the implementation of virtual technologies and strategies to continue with its academic programs.^1^ This opened a great opportunity to discover and innovate in virtual education. One of these innovative tools for virtual education are the massive open online courses (MOOCs) which are often free of charge^2^ and can reach massive, as its name implies, amounts of people, the most popular MOOC had reached more than 2.5 millions of people for very diverse backgrounds.^3^ This technology had also been implemented in formal medical education curriculums^4–6^ and even the world health organization (WHO) had taken advantage of this technology to give knowledge and reach a vast amount of healthcare professionals and students in times of the new coronavirus disease of 2019^7^ (COVID-19). These video based courses are not only specially relevant and useful in this time where quarantines and virtual education had become the new normality for many medical students but also had been proved to be equally effective when compared to live lecture courses.^8^

The International League against epilepsy (ILAE) in 2014 created the epilepsy education task force with the objective to create a web-based virtual campus to teach about epilepsy on a competence-based approach, this goal has not been reached yet but they had published what they consider should be the learning domains that should be teach in epilepsy^9^ for healthcare professionals involved in the care of people with epilepsy. Using these domains proposed by the ILAE a MOOC on epilepsy was made for Latin American healthcare students during the COVID-19 pandemic.

## MATERIALS OR PATIENTS AND METHODS

### The MOOC

A Seven week MOOC about the principles on diagnosis and management of epilepsy was made using free platforms and software. The MOOC was divided into 5 main topics, see **Table 1**, based on a competence-based curriculum in epileptology published previously by the ILAE.^9^ Each lecture consisted of a topic review presented with PowerPoint slides with the lecturer voice-over and freehand drawings using a digital tablet, see **Figure 1**. Each lecture was presented via YouTube live and the recorded lecture was then upload to the Google classroom were the MOOC was hosted. The screen recording and broadcast through YouTube was made using Open Broadcaster Software, an open and free software. In Google classroom during the seven weeks the MOOC lasted, all the students had access to the lectures recordings, the scientific articles used in the topic review a forum and a five question review test for each lecture; the tests were made using Google forms. All the previously mentioned Google products are free to use.

**Table 1.** Epilepsy MOOC schedule.

| epilepsy MOOC schedule |  |  |
| --- | --- | --- |
| Main topic | secondary topics | Lenght (mm:ss) |
| Pathophysiology of epilepsy | Epileptogenesis; Ictogenesis | 25:24 |
| non-epileptic paroxysmal events | Psychogenic Nonepileptic Seizures | 49:55 |
| Principles of electroencephalography | Differential amplification; common EEG montages; Normal EEG patterns; Epileptiform discharges characteristics | 74:10 |
| ILAE 2017 seizure classification | Diagnostic levels of epilepsy | 61:15 |
| pharmacologic principles of epilepsy treatment | Action potentials; Blood brain barrier; Pharmacological and pharmacodynamic review of antiepileptic drugs | 52:11 |

**Figure 1.**
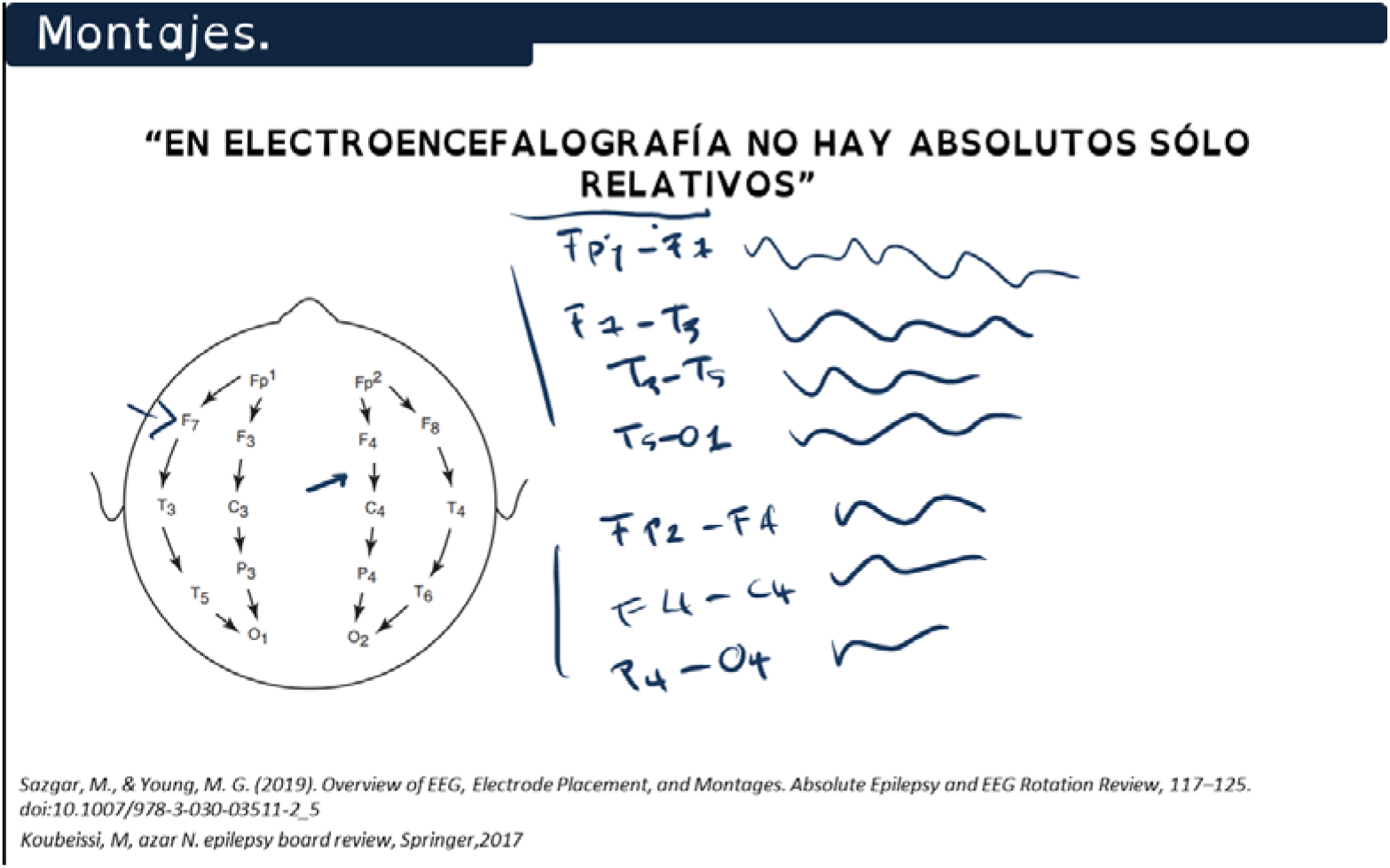
Lecturer drawing over a PowerPoint slide used in the MOOC also known as the khan-style lecture production.

### The participants

The MOOC was promoted trough the social networks Whatsapp and Instagram all the participants sign up through a Google Form were a record of their personal data was made. Using the E-mails recorded with the forms they were enrolled in the Google classroom where the MOOC were hosted. After the last lecture the participants were invited to answer a similar form in order to compare the data from the cohort who signed up for the MOOC and those who finished it. In the forms the participants were asked for their permission for the use of their personal data and only the personal data of the participants which agreed is presented in this manuscript.

## RESULTS

A Total of 250 healthcare students signed up in the epilepsy MOOC, the demographic characteristics of the participants are summarized in the **table 2**, from a total of 8 different Latin American countries, see **Figure 2**. From these 250 participants only 43 (17.2%) of them had previously participated in other MOOCs. After the completion of the MOOC, when asked about the length of the lectures of the MOOC 94% of the participants who finished the MOOC said they had an optimal Length, see **Figure 3**, and 98.3% of them agreed that they would participate in future MOOCs in the future. This epilepsy MOOC had a completion rate of 37.2% for a total of 93 participants.

**Table 2.**
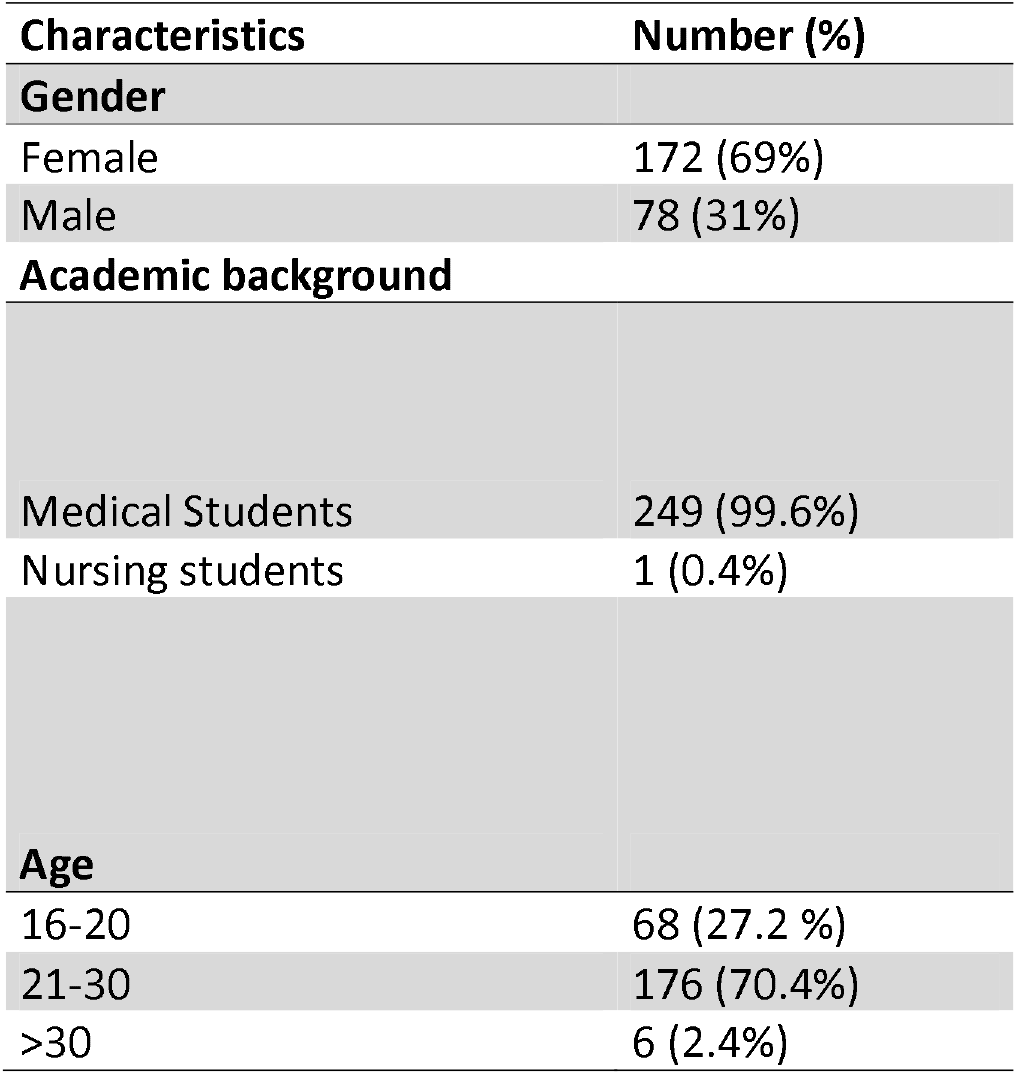
Characteristics of the students which signed up for the MOOC.

| Characteristics | Number (%) |
| --- | --- |
| **Gender** |  |
| Female | 172 (69%) |
| Male | 78 (31%) |
| **Academic background** |  |
| Medical Students | 249 (99.6%) |
| Nursing students | 1 (0.4%) |
| **Age** |  |
| 16-20 | 68 (27.2 %) |
| 21-30 | 176 (70.4%) |
| >30 | 6 (2.4%) |

**Figure 2.**
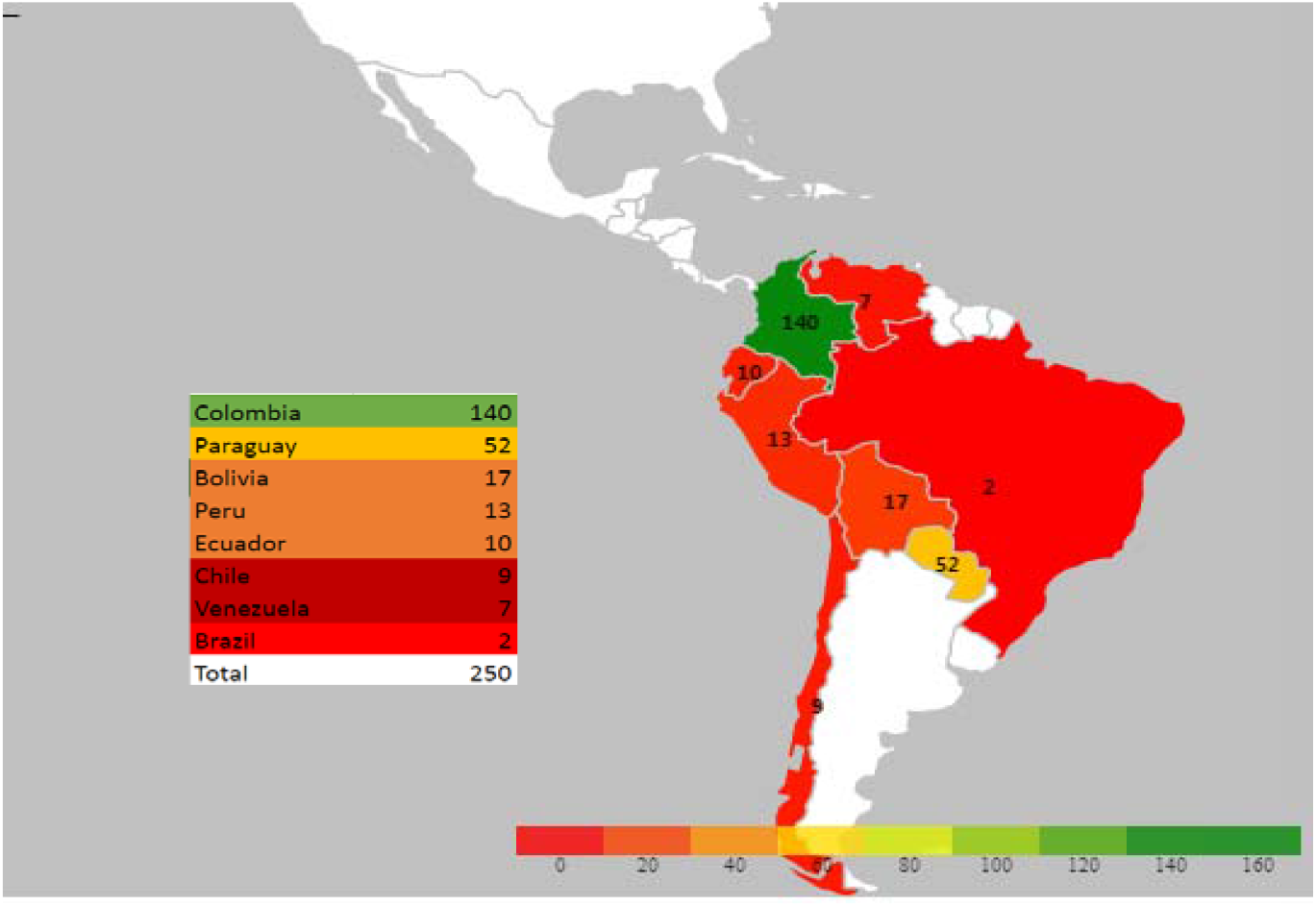
Heat map Showing the countries of origin of the participants of the MOOC.

**Figure 3.**
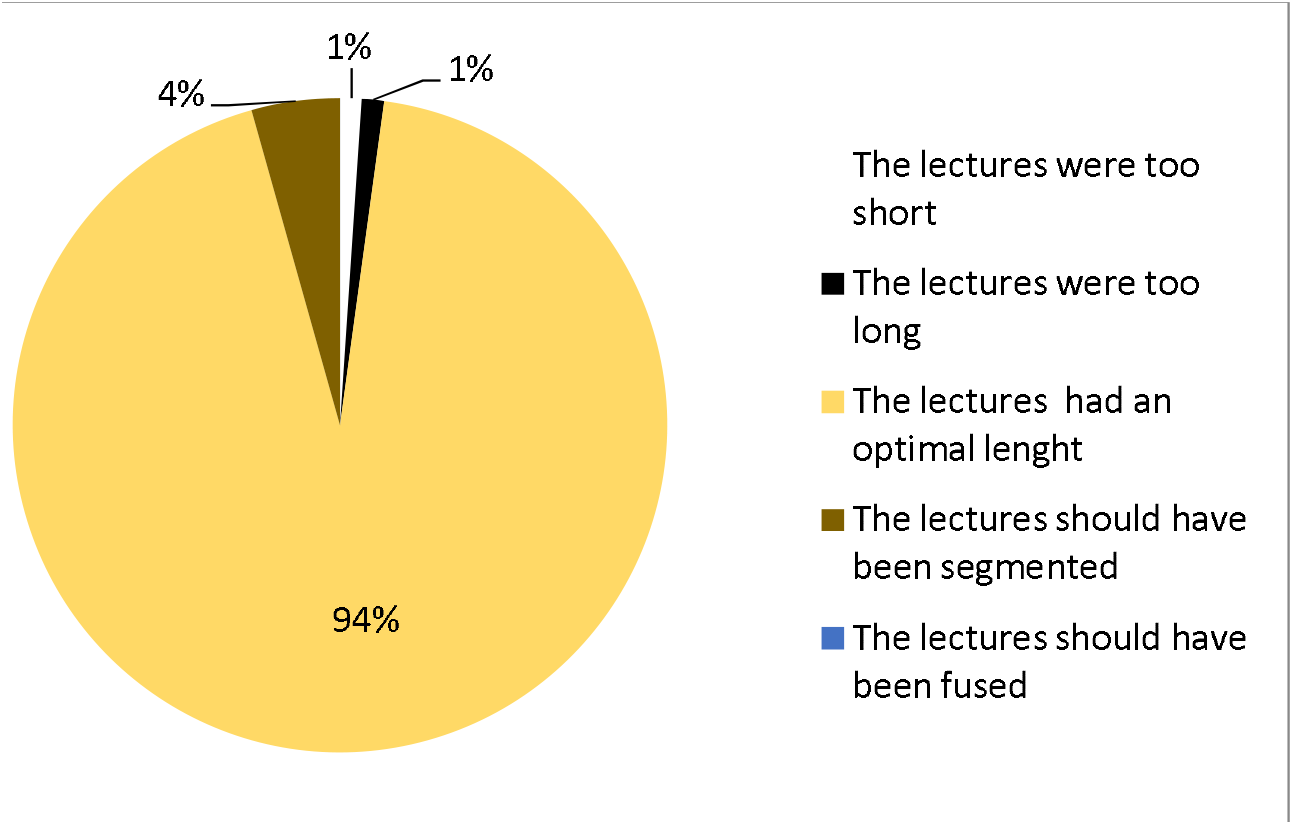
Opinion upon the lecture length from the participants which finished the MOOC.

## DISCUSSION

Student-made content for students are not novel, nevertheless with the implementation of Information and communications technologies this content can reach more than the author’s own cohort and it can break gaps that were unbreakable before. The geographical distance can be shortened and high quality resources can be accessible for almost anyone with an internet connected device.^10^ MOOCs are part of these technologies it was introduced in 2008 as a concept by Cormier and Alexander and it has become an immensely popular tool since then.^11^ This Experience making a peer teach epilepsy MOOC it’s an important statement on how Latin American students which are usually a hard population to reach with conventional MOOC technologies^10,12^ are indeed reachable and open to learn using this tool. One of the main determinants that acts as a barrier to the implementation of MOOCs in Latin American countries is the language barrier, this is perfectly shown in the report of the data from a WHO MOOC on COVID-19 the main source of enrolments were in their Spanish version of the MOOC,^7^ this show how you can reach this population if you break this barrier; this is also shown here, this epilepsy MOOC was given entirely in Spanish as a result only two students from brazil signed up, this two students lived in border cities with Spanish speaking countries so they spoke Spanish as a second language and overcame this barrier.

This MOOC had a completion rate of 37.2% this may seem low at a first glance but MOOCs are known to have low completion rates having a averages of completion rates of less than 10% and as low as 6,5%.^13–14^ Our success may be due to two main factors one of them is that we gave free certificates for those who finished the MOOC and the second one could be the methodology: first the khan-style production style was used in this MOOC, Khan-style consist in a full screen video with the lecturer drawing freehand on a digital tablet, this method had shown more engagement from students compared with other methods of lecture production like having PowerPoint slides only.^15^ The second methodological aspect that may have had a positive impact on the completion rate may be the self-spacing nature of this MOOC, students could revisit any topic at any time as long as it was in the 7 week period the MOOC was open this has shown better results and it’s associated with better learning.^16^ And the third methodological aspect of this MOOC that may had had a positive influence in the completion rate is the length of the lectures; the length of the lectures in this MOOC was in average of 52 minutes this is unconventional for MOOCs and it’s a statement that may contradict previous findings that show otherwise.^15^ However 94% of the students who finished this MOOC affirmed that the lecture length was optimal. This result may be biased because we only use the data of those who finished the MOOC which may be more pleased with the MOOC in comparison with those who dropped from the MOOC without finishing it.

Only the 17.2% of the participants in this MOOC had previously participated in other MOOCs but when asked to the students who finished the course if they would participate in MOOCs again 98.9% of them said they would. This can show us that when breaking the barriers that keep the gaps for Latin American Students to acquire knowledge trough MOOCs we can start building a MOOC-participative culture in them.

In conclusion MOOCs can be easily implemented as a powerful pedagogic strategy during the COVID-19 pandemic and can have a positive impact not only in in its proposed learning objectives but it can help closing the gap that prevent Latin American healthcare students to acquire actively knowledge trough them.

## Data Availability

All the data regarding this article is presented in the manuscript itself.

## Acknowledgment

The author would like to acknowledge Ana María Riveros Betancour for proofreading this manuscript and her invaluable advice and Valentina Parra and Sofia Muñoz two medical students which helped with the logistics for the making of the Massive online open course this paper is based on.

## Financing

The author did not receive any funding for the making of this paper

## Conflict of interest statement by authors

The author declares no conflict of interest

## Compliance with ethical standards

The participants of this study signed a digital informed consent and only the data from the participants who agreed their data to be used is shown in this manuscript.

## Authors Contribution Statement

**Authors Contribution Statement:**
| Contributor Role | Role Definition | Authors |  |  |  |  |  |
| --- | --- | --- | --- | --- | --- | --- | --- |
|  |  | 1 | 2 | 3 | 4 | 5 | 6 |
| **Conceptualization** | Ideas; formulation or evolution of overarching research goals and aims. | X |  |  |  |  |  |
| **Data Curation** | Management activities to annotate (produce metadata), scrub data and maintain research data (including software code, where it is necessary for interpreting the data itself) for initial use and later reuse. | X |  |  |  |  |  |
| **Formal Analysis** | Application of statistical, mathematical, computational, or other formal techniques to analyze or synthesize study data. | X |  |  |  |  |  |
| **Funding Acquisition** | Acquisition of the financial support for the project leading to this publication. |  |  |  |  |  |  |
| **Investigation** | Conducting a research and investigation process, specifically performing the experiments, or data/evidence collection. | X |  |  |  |  |  |
| **Methodology** | Development or design of methodology; creation of models | X |  |  |  |  |  |
| **Project Administration** | Management and coordination responsibility for the research activity planning and execution. | X |  |  |  |  |  |
| **Resources** | Provision of study materials, reagents, materials, patients, laboratory samples, animals, instrumentation, computing resources, or other analysis tools. | X |  |  |  |  |  |
| **Software** | Programming, software development; designing computer programs; implementation of the computer code and supporting algorithms; testing of existing code components. | X |  |  |  |  |  |
| **Supervision** | Oversight and leadership responsibility for the research activity planning and execution, including mentorship external to the core team. | X |  |  |  |  |  |
| **Validation** | Verification, whether as a part of the activity or separate, of the overall replication/reproducibility of results/experiments and other research outputs. |  |  |  |  |  |  |
| **Visualization** | Preparation, creation and/or presentation of the published work, specifically visualization/data presentation. | X |  |  |  |  |  |
| **Writing – Original Draft Preparation** | Creation and/or presentation of the published work, specifically writing the initial draft (including substantive translation). | X |  |  |  |  |  |
| **Writing – Review & Editing** | Preparation, creation and/or presentation of the published work by those from the original research group, specifically critical review, commentary or revision – including pre- or post-publication stages. | X |  |  |  |  |  |

